# Tattooing and risk of melanoma: a population-based case-control study in Utah

**DOI:** 10.1101/2025.03.14.25323969

**Authors:** Rachel D. McCarty, Britton Trabert, Lindsay J. Collin, Morgan M. Millar, David Kriebel, Laurie Grieshober, Mollie E. Barnard, Jenna Sawatzki, Marjorie Carter, Valerie Yoder, Jeffrey A. Gilreath, Douglas Grossman, John Hyngstrom, Paul J. Shami, Jennifer A. Doherty

## Abstract

**Background:** Tattooing can deliver carcinogens directly into the skin and cause immunologic responses, yet the relationship between tattooing and melanoma risk is unknown.

**Methods:** In a population-based case-control study with 1,167 melanoma cases (566 in situ; 601 invasive) and 5,835 frequency-matched controls, we examined tattooing and melanoma risk using multivariable logistic regression to calculate odds ratios (ORs) and 95% confidence intervals (CIs).

**Results:** While ever receiving a tattoo was not strongly associated with melanoma risk, heavier tattooing exposure was associated with decreased risk. Melanoma risk was decreased among individuals who had received four or more tattoo sessions (OR 0·44 [95% CI 0·27–0·67]) and individuals who had three or more large tattoos (OR 0·26 [95% CI 0·10–0·54) compared with those who were never tattooed. Invasive melanoma risk was decreased among individuals who received their first tattoo before age 20 (OR 0·48 [95% CI 0·29–0·82]) compared with never tattooed individuals. These patterns were stronger among men than women. Conversely, individuals who had only received one tattoo session had a higher risk of melanoma (OR 1·53 [95% CI 1·16–2·00) compared with individuals who were never tattooed, particularly for in situ melanoma (OR 1·85 [95% CI 1·31–2·63).

**Conclusions:** Our findings suggest a complex relationship between tattooing and melanoma risk. There was evidence of reduced melanoma risk with more tattoo exposure, but also increased risk among those who were only tattooed once. The potential causes of these seemingly contradictory associations could include a variety of factors including sun exposure-related behaviors or immune responses to timing and quantities of tattooing. These findings justify further investigation.

## Introduction

As of 2023, 38% of women and 27% of men in the United States (U.S.) were estimated to have a tattoo,^1^ but the long-term health effects of tattooing are largely unknown. Many commercially available tattoo inks contain carcinogens including metals, polycyclic aromatic hydrocarbons (PAHs), and primary aromatic amines (PAAs).^2^ In addition, inks can photodegrade when exposed to ultraviolet radiation and form new toxic compounds.^3–5^ Rather than remaining fully in the skin, inks and their carcinogenic components accumulate in regional lymph nodes.^6,7^ Tattooing can also result in long-term inflammatory and immune responses as allergies from tattooing can occur months to years after tattooing.^8,9^ These immune effects may be relevant to melanoma risk as most melanoma tumors are highly mutagenic and elicit complex immune responses.^10^ In case reports, over 160 skin cancer cases have been observed within tattoos, including at least 43 cases of melanoma.^11^ However, only one epidemiologic study of tattooing and melanoma risk has been published to date which observed increased risk of combined melanoma and non-melanoma skin cancer associated with ever getting tattooed and ever receiving a large tattoo.^12^ To our knowledge no published studies to date have examined whether tattooing may increase melanoma risk alone.

In this study, we evaluated associations between tattoo exposures and melanoma incidence in Utah, the state with the highest melanoma incidence in the U.S.^13^ Melanoma risk increases with age, but patterns of melanoma incidence by age vary between women and men.^14^ In the U.S. population, women have a higher risk of melanoma than men prior to age 45 years.^14^ Conversely, after age 45, the risk of melanoma is higher among men than among women.^14^ As patterns of tattooing and melanoma incidence differ considerably between women and men, in addition to overall patterns we examined associations separately by sex.

## Methods

We conducted a population-based case-control study. A flow chart showing the inclusion/exclusion process is shown in Figure 1. All incident cases of in situ and invasive melanoma diagnosed in Utah between January 1, 2020 and June 30, 2021, ages 19–79 years old were identified by the Utah Cancer Registry using rapid case ascertainment and mailed a letter inviting them to participate in the study. A trained interviewer attempted to contact each case via telephone using a standard protocol. Of the 3,032 incident cases, 133 were determined to be eligible outside of the interviewing time frame ending in January 2024 for a total of 2,899 cases included in recruitment. Of the 2,899 cases, 105 (4%) were deceased before first contact, 1,111 (38%) were unable to be reached, and 505 (17%) refused. Telephone surveys were completed by 1,178 individuals for a response proportion of 41%. Individuals were excluded after interview if they were not a Utah resident one year prior to dagnosis (n=5), or were missing tattoo data (n=6). The final analytic dataset included 1,167 melanoma cases (566 in situ and 601 invasive).

**Figure 1.**
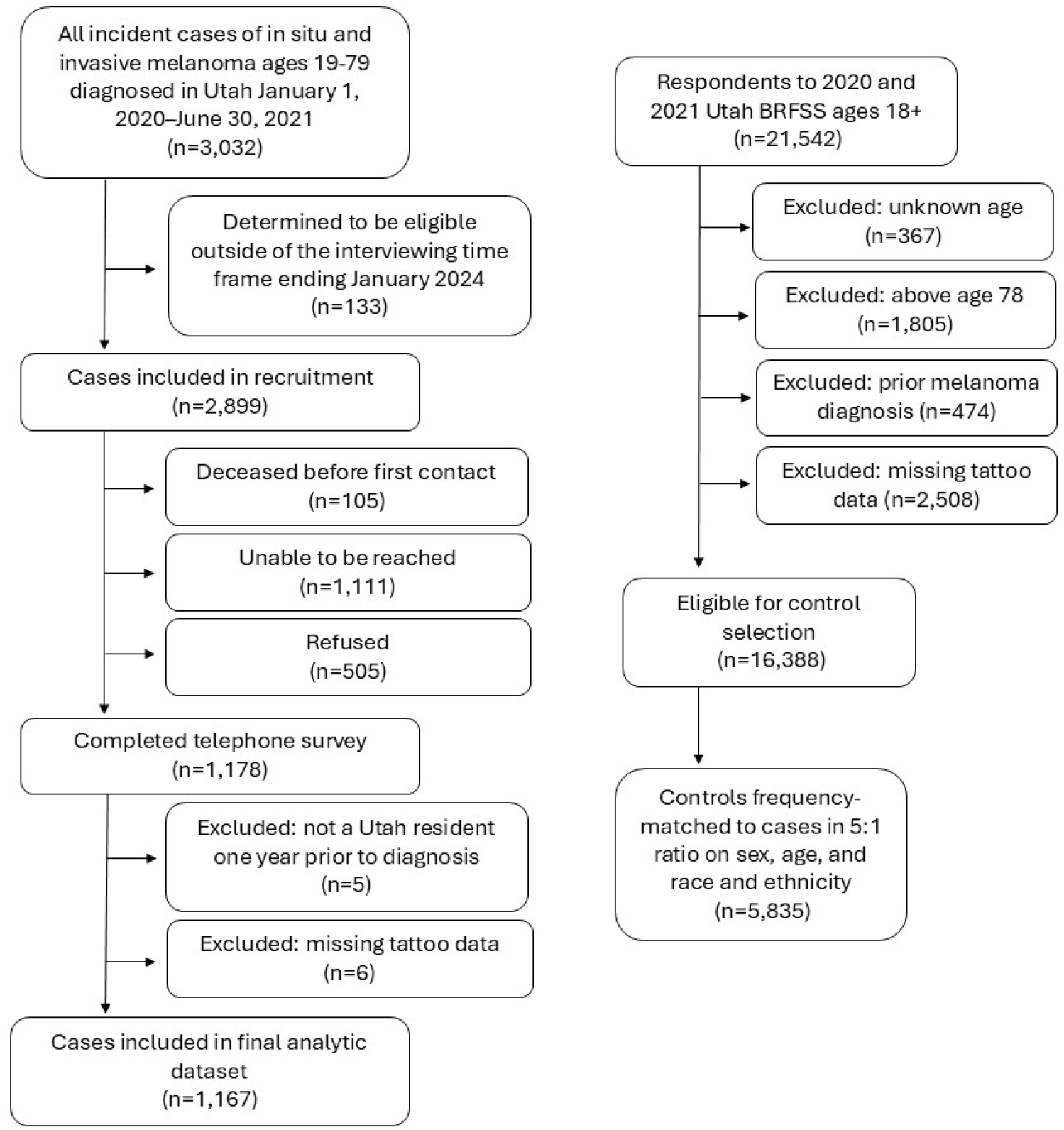
Flow chart of included melanoma cases and frequency-matched controls in Utah Abbreviation: Behavioral Risk Factor Surveillance System (BRFSS)

Controls were selected from respondents to the 2020– 2021 Utah Behavioral Risk Factor Surveillance System (BRFSS) survey. As previously described,^15^ we worked with the Utah Department of Health and Human Services to add three tattoo questions to the BRFSS survey: 1) What is the total number of tattooing sessions you have had? 2) How many of your tattoos are bigger than your palm? and 3) How old were you when you got your first tattoo? Participants were asked to include every tattoo they had ever received that was administered using a tattoo machine. Surveys were completed by 21,542 participants with response proportions of 55% in 2020 and 47% in 2021. We excluded individuals with unknown age (n=367), individuals above age 78 years (n=1,805) to be comparable to cases who were ages 19–79, as cases were asked about the time period one year prior to diagnosis, individuals who reported a prior melanoma diagnosis (n=474), and individuals who were missing tattoo data (n=2,508), leaving 16,388 participants eligible for control selection. We frequency-matched controls to cases in a 5:1 ratio on sex, five-year age group, and race and ethnicity (Hispanic, non-Hispanic (NH) American Indian or Alaska Native, NH Asian, NH Black, NH Pacific Islander, NH White, NH multiracial, NH other, unknown). In total, 5,835 controls were selected.

### Statistical analysis

We computed odds ratios (ORs) and 95% confidence intervals (CIs) from logistic regression models associating tattoo exposures (ever tattooed; time since first tattoo; age at first tattoo; number of tattoo sessions; number of large tattoos) with any melanoma diagnosis, and separately for in situ and invasive melanoma. Models were adjusted for sex, age (five-year groups), race and ethnicity (NH White, Hispanic, all other racial and ethnic groups), education (<high school diploma, high school diploma, some college, four-year college degree or more), ever smoking (yes/no), physical activity in the past 30 days (yes/no), and body mass index (BMI; <25/25+ kg/m^2^). We then fit models stratified by sex.

Data on the melanoma risk factors indoor tanning, sunscreen use on a sunny summer day, having had at least one red or painful sunburn in the 1–2 year period prior to diagnosis, tattoo sun exposure in the summer, ability to tan, hair and eye color, and personal and family history of melanoma were collected for cases but were not available for controls because of limits on the number of questions that could be added to the BRFSS. In supplemental analyses we computed frequencies of these factors in cases, among those who never received a tattoo, those with one tattoo session, and those with two or more tattoo sessions.

We used propensity score models to explore potential confounders in the relationships between ever tattooing and number of tattoo sessions with melanoma risk.^16^ We first constructed models separately to predict ever receiving a tattoo, one tattoo session, and two or more tattoo sessions compared to never receiving a tattoo using demographic, health, and risk-taking variables within the BRFSS control population. We took an iterative approach by dropping variables, collapsing categories, and considering two-way interactions while utilizing the Akaike information criterion (AIC) to select the optimal model. We then used comparable models including all variables available among cases and controls to match exposed and nonexposed individuals in a 10:1 ratio. We used nearest neighbor matching with a caliper of 0.2 to improve covariate balance using the MatchIt package in R.^17^ We fit logistic regression models to calculate ORs and 95% CIs within the propensity score-matched sets associating tattoo exposures with melanoma risk.

As this study took place in Utah where over half of the population identifies as members of the Church of Jesus Christ of Latter-day Saints (LDS) and patterns of lifestyle variables such as tobacco use differ between LDS and non-LDS individuals,^18^ we included LDS affiliation as a variable in propensity score models. Because LDS affiliation has a strong negative correlation with tobacco use, LDS affiliation was not included in the main multivariable models to avoid collinearity with ever smoking. All analyses were conducted using R Statistical Software (v4·3·1; R core team 2023; Vienna, Austria).

## Results

### Study participants

Cases tended to have a higher education level, were less likely to smoke, and were more likely to engage in physical activity in the last 30 days than controls (Table 1). Men tended to be older at the time of diagnosis than women. BMI was balanced between cases and controls in men.

**Table 1.** Demographic characteristics of in situ and invasive melanoma cases and controls, by sex.

|  | Men |  |  |  |  |  | Women |  |  |  |  |  |
| --- | --- | --- | --- | --- | --- | --- | --- | --- | --- | --- | --- | --- |
|  | Controls |  | In situ cases |  | Invasive cases |  | Controls |  | In situ cases |  | Invasive cases |  |
|  | (n=3,202) |  | (n=320) |  | (n=321) |  | (n=2,633) |  | (n=246) |  | (n=280) |  |
|  | n | (%) | n | (%) | n | (%) | n | (%) | n | (%) | n | (%) |
| **Age** |  |  |  |  |  |  |  |  |  |  |  |  |
| <45 | 464 | (14%) | 44 | (14%) | 49 | (15%) | 689 | (26%) | 65 | (26%) | 72 | (26%) |
| 45-54 | 349 | (11%) | 36 | (11%) | 33 | (10%) | 477 | (18%) | 42 | (17%) | 55 | (20%) |
| 55-64 | 826 | (26%) | 83 | (26%) | 82 | (26%) | 707 | (27%) | 77 | (31%) | 64 | (23%) |
| 65-74 | 1213 | (38%) | 125 | (39%) | 118 | (37%) | 615 | (23%) | 47 | (19%) | 76 | (27%) |
| 75+ | 350 | (11%) | 32 | (10%) | 39 | (12%) | 145 | (6%) | 15 | (6%) | 13 | (5%) |
| **Race and ethnicity** |  |  |  |  |  |  |  |  |  |  |  |  |
| Hispanic | 20 | (1%) | * | (<5%) | * | (<5%) | 30 | (1%) | * | (<5%) | * | (<5%) |
| NH American Indian/Alaskan Native | 0 | (0%) | 0 | (0%) | 0 | (0%) | 0 | (0%) | 0 | (0%) | 0 | (0%) |
| NH Asian | * | (<1%) | * | (<1%) | 0 | (0%) | 15 | (1%) | * | (0%) | * | (<5%) |
| NH Black | * | (<1%) | 0 | (0%) | * | (<1%) | 0 | (0%) | 0 | (0%) | 0 | (0%) |
| NH Pacific Islander | 0 | (0%) | 0 | (0%) | 0 | (0%) | 0 | (0%) | 0 | (0%) | 0 | (0%) |
| NH White | 3110 | (97%) | 311 | (97%) | 311 | (97%) | 2545 | (97%) | 236 | (96%) | 273 | (98%) |
| NH multiracial | 42 | (1%) | * | (<5%) | * | (<5%) | 33 | (1%) | * | (<5%) | * | (<5%) |
| NH Other | * | (<1%) | 0 | (0%) | * | (<5%) | * | (<1%) | 0 | (0%) | 0 | (0%) |
| Missing | 15 | (<1%) | * | (<1%) | * | (<5%) | * | (<1%) | * | (0%) | 0 | (0%) |
| **Education** |  |  |  |  |  |  |  |  |  |  |  |  |
| Less than high school diploma | * | (<5%) | * | (1%) | * | (<5%) | * | (<5%) | * | (<1%) | * | (<1%) |
| High school diploma/GED | 563 | (18%) | 43 | (13%) | 50 | (16%) | 545 | (21%) | 39 | (16%) | 60 | (21%) |
| Some college | 981 | (31%) | 81 | (25%) | 87 | (27%) | 989 | (38%) | 72 | (29%) | 91 | (32%) |
| College graduate or more | 1561 | (49%) | 191 | (60%) | 180 | (56%) | 1030 | (39%) | 134 | (54%) | 127 | (45%) |
| Missing | * | (<1%) | * | (<1%) | * | (<1%) | * | (<1%) | * | (<1%) | * | (<1%) |
| **Ever smoking** |  |  |  |  |  |  |  |  |  |  |  |  |
| Yes | 975 | (30%) | * | (26%) | 88 | (27%) | * | (<25%) | * | (19%) | * | (<15%) |
| No | 2213 | (69%) | 235 | (73%) | 233 | (73%) | 2076 | (79%) | 198 | (80%) | 239 | (85%) |
| Missing | 14 | (<1%) | * | (<1%) | 0 | (0%) | * | (<1%) | * | (<1%) | * | (<1%) |
| **Physical activity in past 30 days** |  |  |  |  |  |  |  |  |  |  |  |  |
| Yes | 2693 | (84%) | 283 | (88%) | 289 | (90%) | 2150 | (82%) | 224 | (91%) | 254 | (91%) |
| No | * | (16%) | 37 | (12%) | * | (<15%) | * | (18%) | 22 | (9%) | * | (<10%) |
| Missing | * | (<1%) | 0 | (0%) | * | (<1%) | * | (<1%) | 0 | (0%) | * | (<1%) |
| **Body mass index** |  |  |  |  |  |  |  |  |  |  |  |  |
| <25 | 726 | (23%) | * | (24%) | * | (<25%) | 851 | (32%) | 123 | (50%) | * | (41%) |
| 25+ | 2404 | (75%) | 240 | (75%) | 243 | (76%) | 1476 | (56%) | * | (47%) | 154 | (55%) |
| Missing | 72 | (<5%) | * | (<5%) | * | (<5%) | 306 | (12%) | * | (<5%) | * | (<5%) |
| **Church of Jesus Christ of Latter-day Saints (LDS) Religion** |  |  |  |  |  |  |  |  |  |  |  |  |
| Yes | 2041 | (64%) | 210 | (66%) | 192 | (60%) | 1640 | (62%) | 145 | (59%) | 171 | (61%) |
| No | 1126 | (35%) | * | (32%) | * | (<40%) | 957 | (36%) | * | (40%) | * | (<40%) |
| Missing | 35 | (<5%) | * | (<5%) | * | (<5%) | 36 | (<5%) | * | (<5%) | * | (<5%) |

\*Censored due to cell values <11
Note: Percentages indicated as "<" are not precise to prevent identification of study participants and may cause columns to not sum to 100%. Race and ethnicity groups have been collapsed in this table to further protect study participant anonymity.
Abbreviation: non-Hispanic (NH)

Among women, a high proportion of controls did not provide their weight resulting in missing values for 12%. Though this reduced the ability to compare patterns between cases and controls, a higher proportion of in situ and invasive cases among women had a BMI <25 compared with controls. The proportion of men and women identifying as LDS was similar between cases and controls.

### Patterns of associations between tattooing exposures and melanoma risk overall

The prevalence of tattooing was 12% among cases and 15% among controls (Table 2). Ever versus never receiving a tattoo was not associated with melanoma risk (melanoma overall, OR 0·92 [95% CI 0·74–1·13]; in situ, OR 1·04 [95% CI 0·78–1·39]; and invasive, OR 0·81 [95% CI 0·60–1·09]). Patterns differed when examining associations of amount of tattoo exposure with melanoma risk. Melanoma risk was decreased among those who received four or more tattoo sessions (melanoma overall, OR 0·44 [95% CI 0·27–0·67]) and among those with three or more large tattoos (melanoma overall, OR 0·26 [95% CI 0·10–0·54]) compared with never receiving a tattoo. These patterns were consistent for both in situ and invasive melanoma. Receiving only one tattoo session was associated with an increased risk of overall melanoma (OR 1·53 [95% CI 1·16–2·00]), and was stronger for in situ (OR 1·85 [95% CI 1·31–2·63]) than invasive melanoma (OR 1·25 [95% CI 0·85–1·83]), compared with never receiving a tattoo. Receiving a first tattoo before age 20 was associated with a decreased risk of invasive (OR 0·48 [95% CI 0·29–0·82]) but not in situ melanoma (OR 0·94 [95% CI 0·60–1·48]) compared with never getting tattooed.

**Table 2.**
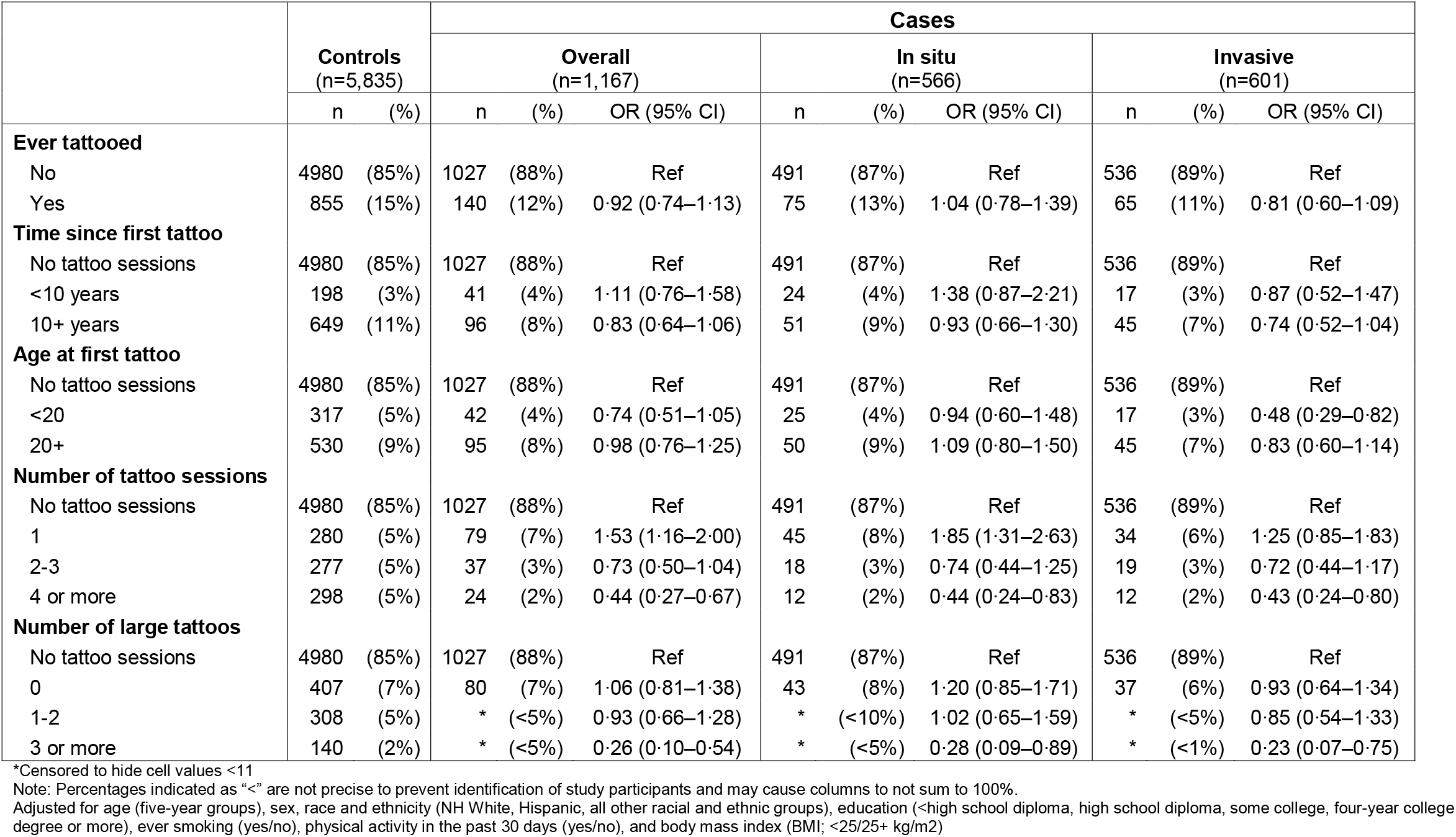
Tattooing exposures and risk of overall, in situ, and invasive melanoma in individuals ages 19-79 years.

### Patterns of associations between tattooing exposures and melanoma risk stratified by sex

The prevalence of tattooing was lower among men (8% among cases and 12% among controls) compared with women (17% among cases and 18% among controls) (Table 3). Ever receiving a tattoo was associated with a decreased risk of melanoma among men (overall melanoma, OR 0·74 [95% CI 0·53–1·02]), but not among women (overall melanoma, OR 1·12 [95% CI 0·84– 1·48]). The decreased melanoma risk associated with four or more tattoo sessions was stronger among men (overall melanoma, OR 0·25 [95% CI 0·09–0·53]; in situ, OR 0·36 [95% CI 0·13– 1·02]; invasive, OR 0·15 [95% CI 0·04–0·62]), and less pronounced among women (overall melanoma, OR 0·64 [95% CI 0·36–1·07]; in situ, OR 0·52 [95% CI 0·23–1·18]; invasive, OR 0·74 [95% CI 0·37–1·48]). The increased melanoma risk associated with one tattoo session was much stronger among women (overall melanoma, OR 1·90 [95% CI 1·30–2·74]; in situ, OR 2·47 [95% CI 1·55–3·93]; invasive, 1·41 [95% CI 0·83–2·39]), than men (overall melanoma, OR 1·21 [0·79–1·81]; in situ, OR 1·31 [95% CI 0·76–2·27]; invasive, OR 1·12 [95% CI 0·64–1·97]).

**Table 3.**
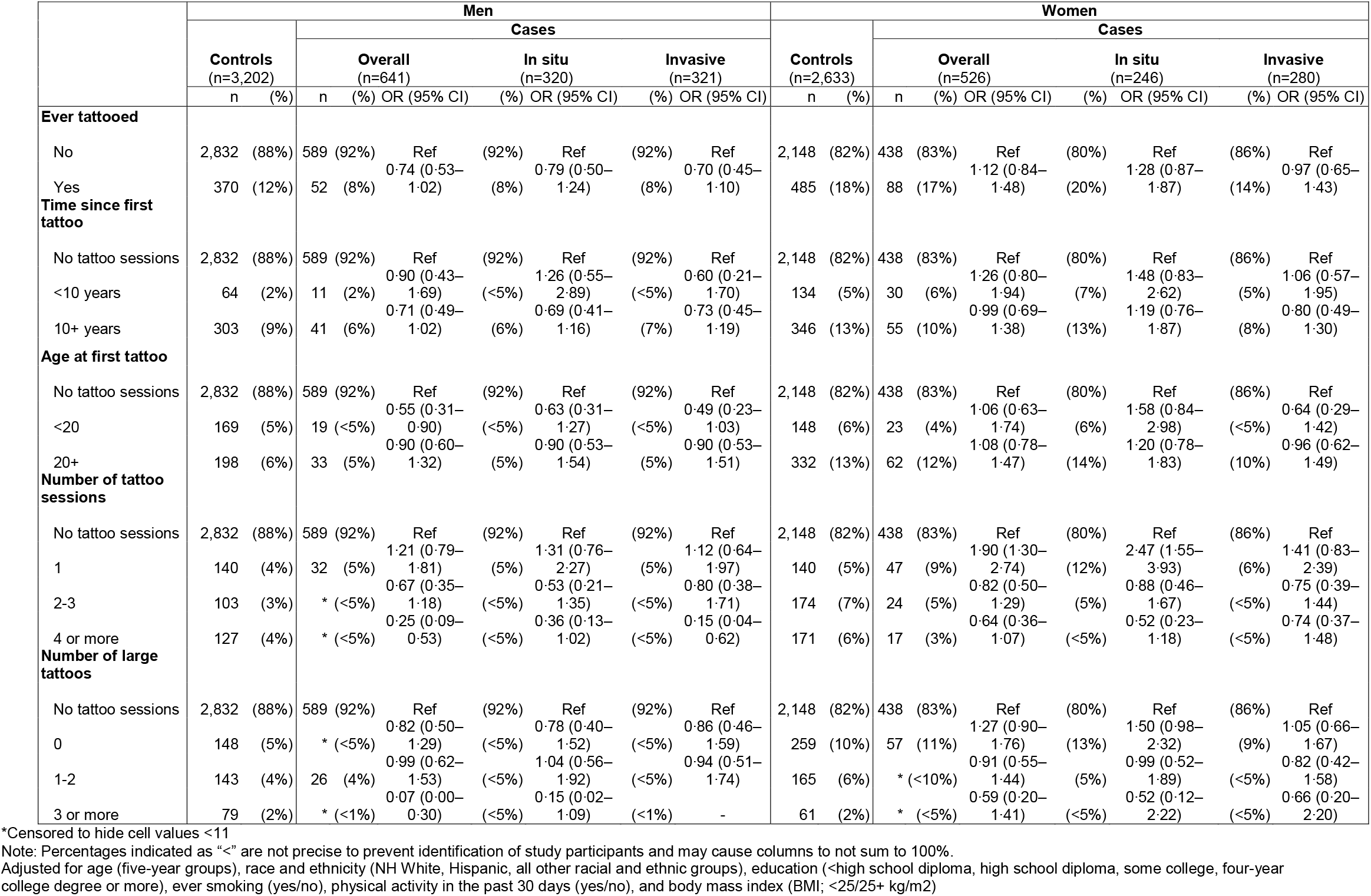
Tattooing exposures and risk of overall, in situ, and invasive melanoma in men and women ages 19–79 years.

Receiving a first tattoo before age 20 was associated with decreased melanoma risk among men for overall (OR 0·55 [95% CI 0·31–0·90]), in situ (OR 0·63 [95% CI 0·31–1·27]), and invasive melanoma (OR 0·49 [95% CI 0·23–1·03]). Among women, receiving a first tattoo before age 20 was associated with decreased risk of invasive (OR 0·64 [95% CI 0·29–1·42]), but not overall (OR 1·06 [95% CI 0·63–1·74]), or in situ melanoma (OR 1·58 [95% CI 0·84–2·98]).

### Distribution of melanoma risk factors among cases and assessment of unmeasured confounding

Behaviors associated with increased melanoma risk tended to be associated with being tattooed among cases. In particular, a higher proportion of tattooed cases had ever used an indoor tanning bed; <40% of men and 69% of women with two or more tattoo sessions, and <30% of men and 53% of women with one tattoo session reported using a tanning device 10 or more times compared with 19% of men and 39% of women with no tattoo sessions (Supplemental Table 1). The proportion of men and women who reported at least one red or painful sunburn in the 1–2 year period prior to diagnosis was highest among those with one tattoo session (53% among men, 51% among women) compared with those with no tattoo sessions (41% among men, 38% among women) and those with 2 or more sessions (<40% among men, 47% among women).

We did not see any major differences between frequency-matched and propensity score-matched results by number of tattoo sessions suggesting there are likely not strong differences in the potential confounders for one tattoo session versus multiple tattoo sessions (Supplemental Table 2).

## Discussion

While the results of this study suggest potentially important roles for tattooing in the incidence of melanoma, as one of the first epidemiologic studies of tattooing and melanoma risk, the findings should be considered as generating hypotheses for further study. The data indicate the possibility of both increased and decreased melanoma risk based on models using variables measuring different aspects of tattooing behavior. Increased melanoma risk was only observed within the lowest level of exposure (one tattoo session) while decreased risk was observed within higher levels of exposure, both for numbers of sessions and numbers of large tattoos.

We initially hypothesized that tattooing may be associated with increased melanoma risk through exposure to carcinogens and inflammatory and immune responses, but the patterns of decreasing risk associated with higher levels of tattoo exposure do not support this hypothesis. It is possible that tattooing may elicit beneficial immune responses that could contribute to immune surveillance of pre-cancerous cells, preventing progression to melanoma.^19^ Additionally, large tattoos could block UV radiation, a major risk factor for melanoma.

While the relationships between melanoma risk factors and tattoos are not fully characterized, a prior study reported that tattooing was associated with a history of painful sunburns, ever use of tanning devices, and lower likelihood of having blue eyes among cancer-free individuals.^20^ In another study from Kluger et al. a higher proportion of tattooed individuals reported both heavier sun exposure and greater use of sun protection compared with non-tattooed individuals.^21^ They also observed individuals with multiple tattoos used sunscreen more frequently and used sunscreens with a higher sun protection factor (SPF) compared with individuals with one tattoo who conversely were more likely to use long sleeves and parasols as sun protection than those with multiple tattoos.^21^ The potential impacts of unmeasured confounding are a consideration for this study as we did not have data on melanoma risk factors among controls, in particular the known melanoma risk factors tanning bed use, sun exposure, family history of melanoma, ability to tan/ sunburn, prior sunburns, and hair and eye color, though we did have this information on cases. In our study, we observed that melanoma cases with tattoos were more likely to use tanning beds, and that ever use of tanning beds increased with higher number of tattoo sessions. We initially anticipated this could be an issue if we observed consistent increased risks of melanoma associated with tattooing. That is, if we observed an increased risk of melanoma associated with tattooing, it could be caused by higher tanning device use among tattooed compared with non-tattooed individuals. However, unmeasured confounding does not appear to be the cause of the *decreased* risk of melanoma we actually observed. For confounding to cause decreased risk estimates, a variable would need to be associated with both increased tattooing and decreased melanoma risk. If anything, unmeasured confounding by tanning bed use may attenuate the decreased risks we observed. It is possible that the *increased* risk of melanoma that we observed with one tattoo session could be due to unmeasured confounding since a higher proportion of cases with one tattoo session reported at least one sunburn in the 1–2 years prior to melanoma diagnosis compared with those who were never tattooed or those with two or more tattoo sessions.

We further examined the potential impacts of unmeasured confounding through propensity score analyses to examine possible confounders in the relationships between ever tattooing and number of tattoo sessions with melanoma risk. The similarities in the propensity score models regardless of number of tattoo sessions and the similarities between the estimates from the original models and the estimates from the propensity score-matched models suggest that unmeasured confounding by factors associated with the available health and demographic variables is not likely to be the cause of the associations we observed.

The existing literature on tattooing and skin cancer is extremely limited. A recent study from Clemmensen et al. composed of two twin-based studies in Denmark reported increased risk of combined melanoma and non-melanoma skin cancers associated with ever receiving a tattoo (case-cotwin hazard ratio (HR) 1·62 [95% CI 1·08–2·41]; cohort HR 3·91 [95% CI 1·42–10.8]) and ever receiving a large tattoo (case-cotwin HR 2·37 [95% 1·11–5·06]).^12^ Within their cohort study, they also reported increased risk of basal cell carcinoma associated with ever tattooing (HR 2·83 [95% CI 1·42–10·78).^12^ The study did not report number of tattoo sessions, but if a large proportion of participants had only one tattoo session, our findings would be consistent with theirs. Notably, Clemensen et al.’s study only included a very small number of individuals with tattoos (8–30 tattooed cancer cases depending on the study design).^12^ Another study by Barton et al. observed a decreased prevalence of tattooing among individuals with early onset basal cell carcinoma compared with cancer free controls.^20^ In a letter to the editor which presented a re-analysis of the data, there was a 40% decreased risk of basal cell cancer among individuals who were ever tattooed (crude OR and 95% CI of 0·6 [0·5–0·7]).^22^ Barton et al. also reported an 80% increased risk of basal cell carcinomas co-occurring in the region of the body that was tattooed.^20^ We were not able to consider tattoo location in our analyses because we did not have information on tattoo location among controls.

The population-based study design using SEER registry data was a strength as it ensured complete ascertainment of all histologically-confirmed melanoma cases in Utah during the study period. Further, the use of BRFSS data ensured controls were reflective of the overall population from which cases arose. Because controls were selected from the BRFSS, we were limited to the variables included on the BRFSS, and a key limitation of this study was the lack of data on potential confounders among controls. We also were not able to ask controls detailed questions on tattooing that may be relevant to melanoma risk including tattoo location and tattoo sun exposure. We were further limited by the 41% response proportion among cases, though this response proportion is typical for interview-based studies.^23^ Despite these limitations, this study provides some of the first evidence to begin understanding how tattooing may relate to melanoma risk.

## Conclusions

Our findings suggest there may be contributions of tattooing to melanoma risk, but further studies are needed. We found some evidence that low exposure to tattooing through one tattoo session is associated with an increased risk of melanoma, but higher exposure such as more tattoo sessions or more large tattoos is associated with decreased risk. We also observed a decreased risk of invasive melanoma associated with receiving a first tattoo before the age of 20 years. The biologic mechanisms underlying these associations are not known but could be related to tumor immune surveillance or reduced UV exposure through tattoo skin coverage. It is also possible that sun exposure-related behaviors influenced some of our results, in particular the increased melanoma risk observed with one tattoo session. Future larger studies are needed to clarify the relationships between tattooing and melanoma risk.

## Data Availability

The data from cancer cases used in this study were obtained from the Utah Cancer Registry which collects and maintains confidential data in accordance with Utah law. These data are not publicly available but can be accessed through review and approval of a proposed study protocol by the University of Utah Institutional Review Board and the Resource for Genetic and Epidemiologic Research. Data from Utah BRFSS participants which were utilized as controls in this study are restricted and maintained by the Utah Department of Health and Human Services, through which these data can be requested.

