## Supplemental Tables 1 and 2 for "Tattooing and risk of melanoma: a population-based case-control study in Utah"

**Supplementary Materials**

**Table of Contents**

Supplemental Table 1	2

Supplemental Table 2	3

**Supplemental Table 1. Distribution of melanoma risk factors among melanoma cases with no tattoos, one tattoo session, and two or more tattoo sessions, by sex**

|  | **Men** | | | **Women** | | |
| --- | --- | --- | --- | --- | --- | --- |
|  | **Never tattooed** (n=590) | **One tattoo session** (n=32) | **2+ tattoo sessions** (n=20) | **Never tattooed** (n=440) | **One tattoo session** (n=47) | **2+ tattoo sessions** (n=42) |
|  | % | % | % | % | % | % |
| **Indoor tanning device use (ever)** |  |  |  |  |  |  |
| None | 62% | 47% | <43% | 37% | <25% | <14% |
| <10 times | <19% | <26% | <21% | <25% | <25% | <19% |
| 10+ times | 19% | <30% | <40% | 39% | 53% | 69% |
| Missing | <1% | <1% | <6% | <1% | <1% | <5% |
| **Continued use of indoor tanning device after first tattoo** |  |  |  |  |  |  |
| Never tanning | ·· | 47% | <42% | ·· | <25% | <15% |
| No | ·· | <33% | <38% | ·· | 49% | 57% |
| Yes | ·· | <23% | <21% | ·· | 26% | 26% |
| Missing | ·· | <1% | <6% | ·· | <5% | <6% |
| **During the summer, are any of your tattoos exposed to the sun when you go outside?** |  |  |  |  |  |  |
| No | ·· | 47% | <35% | ·· | 45% | 29% |
| Yes | ·· | 53% | <75% | ·· | 55% | 71% |
| **How often do you try to keep your tattoos protected from the sun i.e., sunscreen or clothing?** |  |  |  |  |  |  |
| Always or most of the time | ·· | <25% | <40% | ·· | 58% | 57% |
| Sometimes or rarely | ·· | <55% | <45% | ·· | <30% | <33% |
| Never | ·· | <25% | <25% | ·· | <15% | <15% |
| **Sunscreen use on a sunny summer day when outside for more than one hour*** |  |  |  |  |  |  |
| Always or more than 50% of the time | 35% | 41% | <32% | 52% | 53% | 50% |
| Less than 50% of the time | 45% | 44% | <51% | 35% | 32% | 26% |
| Never | <20% | <18% | <22% | <15% | <10% | <20% |
| Missing | <5% | <1% | <1% | <5% | <10% | <10% |
| **At least one red or painful sunburn in past 12 months**** |  |  |  |  |  |  |
| No | 54% | <35% | 65% | 55% | <50% | <47% |
| Yes | 41% | 53% | <40% | 38% | 51% | 47% |
| Missing | 4% | <18% | <1% | 6% | <1% | <10% |
| **Ability to tan** |  |  |  |  |  |  |
| Tans deeply | 13% | <25% | <27% | 9% | <15% | <25% |
| Tans moderately | 51% | 44% | <42% | 41% | 40% | 40% |
| Tans lightly | 28% | <27% | <36% | 32% | 36% | 26% |
| No tan | <10% | <10% | 0% | 15% | <13% | <13% |
| Missing | <5% | 0% | 0% | 2.5% | 0% | 0% |
| **Hair and eye color** |  |  |  |  |  |  |
| All other hair + eye color combinations | 98% | <97% | <95% | 94% | <95% | 98% |
| Red hair blue eyes | <5% | <10% | <13% | <6% | <6% | <5% |
| Missing | <1% | 0% | 0% | <5% | 0% | <1% |
| **Personal history of melanoma** |  |  |  |  |  |  |
| No | 83% | 97% | 80% | 85% | 79% | 93% |
| Yes | <20% | <5% | <20% | <15% | <20% | <10% |
| Missing | <5% | <1% | <7% | <5% | <5% | <1% |
| **Family history of melanoma** |  |  |  |  |  |  |
| No | 71% | <75% | <62% | 66% | <80% | <80% |
| Yes | <30% | <30% | <41% | 34% | <25% | <25% |
| Missing | <1% | 0% | 0% | 0% | 0% | 0% |

*Asked for the time period one year prior to diagnosis

**Asked for the time period 1–2 years prior to diagnosis

Note: Percentages indicated as “<” are not precise to prevent identification of study participants and may cause columns to not sum to 100%.

**Supplemental Table 2. Results from frequency-matched models and propensity score-matched models for tattoo exposures and overall melanoma risk, separately for ever tattooed, one tattoo session, or two or more tattoo sessions, compared to never tattooed**

|  | Ever/never tattooed | | | One tattoo session/never tattooed | | | 2+ tattoo sessions/never tattooed | |
| --- | --- | --- | --- | --- | --- | --- | --- | --- |
|  | Frequency-matched model (n=1,167) | Propensity score-matched model** (n=623) | Frequency-matched model (n=1,106) | | Propensity score-matched model** (n=462) | Frequency-matched model (n=1,088) | | Propensity score-matched model** (n=417) |
|  | OR (95% CI) | OR (95% CI) | OR (95% CI) | | OR (95% CI) | OR (95% CI) | | OR (95% CI) |
| **Ever tattooed** |  |  |  | |  |  | |  |
| No | Ref | Ref | Ref | | Ref | Ref | | Ref |
| Yes | 0·92 (0·74–1·13) | 0·83 (0·66–1·04) | 1·51 (1·14–1·97) | | 1·36 (1·03–1·77) | 0·60 (0·44–0·80) | | 0·62 (0·44–0·84) |
| **Time since tattoo** |  |  |  | |  |  | |  |
| No tattoo sessions | Ref | Ref | Ref | | Ref | Ref | | Ref |
| <10 years | 1·11 (0·76–1·58) | 1·13 (0·76–1·62) | 1·42 (0·88–2·21) | | 1·52 (0·95–2·33) | 0·82 (0·44–1·41) | | 0·82 (0·44–1·43) |
| 10+ years | 0·83 (0·64–1·06) | 0·72 (0·55–0·94) | 1·53 (1·09–2·12) | | 1·29 (0·92–1·78) | 0·53 (0·37–0·74) | | 0·55 (0·37–0·79) |
| **Age at tattoo** |  |  |  | |  |  | |  |
| No tattoo sessions | Ref | Ref | Ref | | Ref | Ref | | Ref |
| <20 | 0·74 (0·51–1·05) | 0·66 (0·42–1·00) | 1·49 (0·84–2·52) | | 1·40 (0·77–2·35) | 0·53 (0·32–0·82) | | 0·71 (0·41–1·15) |
| 20+ | 0·98 (0·76–1·25) | 0·88 (0·68–1·13) | 1·49 (1·08–2·02) | | 1·36 (0·99–1·82) | 0·62 (0·42–0·89) | | 0·56 (0·37–0·81) |
| **Large tattoos** |  |  |  | |  |  | |  |
| No tattoo sessions | Ref | Ref | Ref | | Ref | Ref | | Ref |
| 0 | 1·06 (0·81–1·38) | 0·99 (0·74–1·29) | 1·33 (0·95–1·81) | | 1·31 (0·94–1·79) | 0·73 (0·46–1·12) | | 0·75 (0·47–1·16) |
| 1–2 | 0·93 (0·66–1·28) | 0·80 (0·55–1·13) | 1·97 (1·16–3·22)* | | 1·41 (0·85–2·21)* | 0·65 (0·42–0·97) | | 0·66 (0·40–1·01) |
| 3 or more | 0·26 (0·10–0·54) | 0·12 (0·02–0·37) | - | | - | 0·26 (0·10–0·56) | | 0·23 (0·07–0·56) |
| **Variables in propensity score-matched model** | Ever tattooed = male + age 18 to 24 + age 25 to 29 + age30 to 34 + age35 to 39 + age40 to 44 + age45 to 49 + age50 to 54 + age55 to 59 + age60 to 64 + age70 to 74 + age75 to 79 + Hispanic + NH Other race + LDS religion + education less than highschool diploma + education highschool diploma + education some college + never tobacco smoking + underweight BMI + obese BMI + married + straight sexuality + no current e-cigarette use + no binge drinking + health insurance + flu shot in past year + never diagnosed with depression + education less than highschool diploma*male + obese*male + married*male + straight sexuality*male + no current e-cigarette use*male + health insurance*male + flu shot in past year*male + never tobacco smoking*LDS religion | | | 1 tattoo session = male + age 18 to 24 + age 25 to 29 + age30 to 34 + age35 to 39 + age40 to 44 + age45 to49 + age50 to 54 + age55 to 59 + age60 to 64 + age70 to 74 + age75 to 79 + Hispanic + NH Other race + LDS religion + *education highschool diploma OR less* + education some college + never tobacco smoking + obese BMI + married + no current e-cigarette use + flu shot in past year + **no marijuana use** + never diagnosed with depression + *education highschool diploma OR less*LDS religion* + never tobacco smoking*LDS religion | | | 2+ tattoo sessions = male + age 18 to 24 + age 25 to 29 + age30 to 34 + age35 to 39 + age40 to 44 + age45 to 49 + age50 to 54 + age55 to 59 + age60 to 64 + age70 to 74 + age75 to 79 + Hispanic + NH Other race + LDS religion + education less than highschool diploma + education highschool diploma **+** education some college + never tobacco smoking + underweight BMI + obese BMI + married + straight sexuality + no current e-cigarette use + no binge drinking + health insurance + flu shot in past year + **no marijuana use** + never diagnosed with depression **+** education less than highschool diploma*male + obese*male + married*male + straight sexuality*male + no current e-cigarette use*male + health insurance*male + flu shot in past year*male | |

*In model for one tattoo session, large tattoos were categorized as 0, or 1 or more, rather than 0, 1–2, and 3 or more.

Abbreviations: body mass index (BMI), Church of Jesus Christ of Latter-day Saints (LDS), non-Hispanic (NH)

** Propensity score models were matched 10:1on the tattooing outcome. As matching was not contingent on case status, the number of cases included in the propensity score matched datasets was smaller than the frequency matched datasets.

Note: Models included ages 19–79.

We selected the best fit propensity score matched model for ever/never tattooed, 1 tattoo session, and 2+ tattoo sessions separately, using a pool of 38 variables. The variables which differed from the model for ever tattooed, but were similar between the models for 1 and 2+ tattoo sessions are **bolded in blue**. The differences in the model for 1 tattoo session which are unique to that model are *italicized in red*.
